# Low dose hydroxychloroquine is associated with lower mortality in COVID-19: a meta-analysis of 26 studies and 44,521 patients

**DOI:** 10.1101/2020.11.01.20223958

**Authors:** Augusto Di Castelnuovo, Simona Costanzo, Antonio Cassone, Roberto Cauda, Giovanni de Gaetano, Licia Iacoviello

**Author notes:** **Address for correspondence** Licia Iacoviello, MD, PhD, Department of Epidemiology and Prevention, IRCCS Neuromed, Via dell’Elettronica, 86077 Pozzilli (IS), Italy.

## Abstract

**Background:** Hydroxychloroquine (HCQ) was proposed as potential treatment for COVID-19, but its association with mortality is not well characterized. We conducted two meta-analyses to evaluate the association between HCQ (with or without azithromycin (AZM)) and total mortality in COVID-19 patients.

**Methods:** Articles were retrieved until October 20^th^, 2020 by searching in seven databases. Data were combined using the general variance-based method on relative risk estimates.

**Results:** A total of 26 articles were found (N=44,521 COVID-19 patients, including N=7,324 from 4 randomized clinical trials (RCTs)); 10 studies were valuable for analysing the association of HCQ+AZM. Overall, the use of HCQ was associated with 21% lower mortality risk (pooled risk ratio: 0.79, 95%CI: 0.67 to 0.93; high level of heterogeneity: I^2^=82%, random effects). This association vanished (1.10, 95%CI: 0.99 to 1.23 and 1.10, 95%CI: 0.99 to 1.23) when daily dose >400 mg or total dose >4,400 mg were used, respectively). HCQ+AZM was also associated with 25% lower mortality risk, but uncertainty was large (95%CI: 0.50 to 1.13; P=0.17). No association was apparent when only pooling the 4 RCTs (13.8% of the overall weight; pooled risk ratio: 1.11, 95%CI: 0.99 to 1.24).

**Conclusions:** HCQ use was not associated with either increased or decreased mortality in COVID-19 patients when 4 RCTs only were evaluated, while a 7% to 33% reduced mortality was observed when observational studies were also included. The association was mainly apparent when pooling studies using lower doses of HCQ. These findings can help disentangling the debate on HCQ use in COVID-19.

**Key-points:** Low dose hydroxychloroquine was associated with reduced mortality in COVID-19 patients, as seen in observational studies but not in randomised clinical trials, which used high doses of hydroxychloroquine. These findings can help disentangling the debate on hydroxychloroquine use in COVID-19.

## INTRODUCTION

The aminoquinoline hydroxychloroquine (HCQ) is an anti-malaria drug, with immunomodulatory and anti-thrombotic properties, currently used in the treatment of autoimmune diseases like rheumatoid arthritis, systemic lupus erythematosus and anti-phospholipid syndrome [1, 2]. At the beginning of the pandemic, it was proposed as a possible therapy in COVID-19 patients since it could directly inhibit viral entry and spread in several *in vitro* and *in vivo* models [3]. Indeed, HCQ has been used in Ebola virus disease [4], human immunodeficiency virus (HIV) infection [5], SARS-CoV-1 infection and the Middle East Respiratory Syndrome [6].

Despite the lack of evidence of efficacy from few randomized clinical trials, HCQ became very popular and widely used by many clinicians. In Italy over 70% of COVID-19 hospitalised patients were treated with HCQ [7]. On the other hand, this drug was given an inexplicable political connotation that shifted the focus more to a political battle than to a discussion based on scientific evidence. The publication of a very questionable study [8] by one of the most reliable scientific journals showing that the use of HCQ was associated to an increased risk of death, lead to the stop of several clinical trials in their tracks and of the HCQ arm in the Recovery trial [9]. This Lancet study was retracted 13 days after publication [10], because its data turned out to be fabricated. Anyway, the principal drug Agencies decided to suspend the authorization to use HCQ for COVID-19. Consequently, the review process of papers on HCQ became increasingly difficult, creating a publication bias that affected all meta-analyses published until now.

However, a number of questions remain open on the relationship between HCQ treatments in COVID-19 patients: is there a dose issue? Does mortality rate of a population or the severity of the disease affect HCQ efficacy? Is there any interaction with other anti-COVID19 drugs? In the last few months, at least three large, well-conducted observational studies have been published showing a protective effect of HCQ on mortality risk in hospitalized COBID-19 patients [7,11,12]. All three studies have not been included in previous meta-analysis [13], and used HCQ doses lower than those administered in randomized clinical trial (RCT), as the Recovery or the Solidarity trials [9,14].

Therefore, we decided to conduct an updated meta-analysis on observational and RCT studies on HCQ use and the mortality outcome in patients hospitalized for COVID-19. We also performed subgroup analyses to dissect whether treatment effects differ according to methodological or clinical characteristics of the primary studies.

## METHODS

This study was conducted according to the recommendations outlined in the Cochrane Handbook for Systematic Reviews of Interventions, version 5.1.0, and reported in line with the PRISMA statement. Institutional review board approval was not required as the study did not directly involve human participants.

### Search strategy

Flow diagram for study selection is reported in Figure 1. Articles published in English were retrieved from inception to October 20^th^, 2020 by searching in Medline, Embase, PubMed, Web of Science, Cochrane Central Database, MedRvix and Preprints.org, with the search terms: “(COVID-19 OR Cov-Sars-2) AND (hydroxychloroquine OR chloroquine)”. In addition, the reference lists of relevant articles for potential studies were also manually reviewed. After initial search, the duplicate results were removed. The remaining articles were screened for relevance by their titles and abstracts by two of us independently (SC and ADC). All selected potential articles were then reviewed by the remaining investigators to ensure their eligibility for inclusion. Disagreements about eligibility of the literature were resolved by consensus based on the agreements of all investigators.

**Figure 1.**
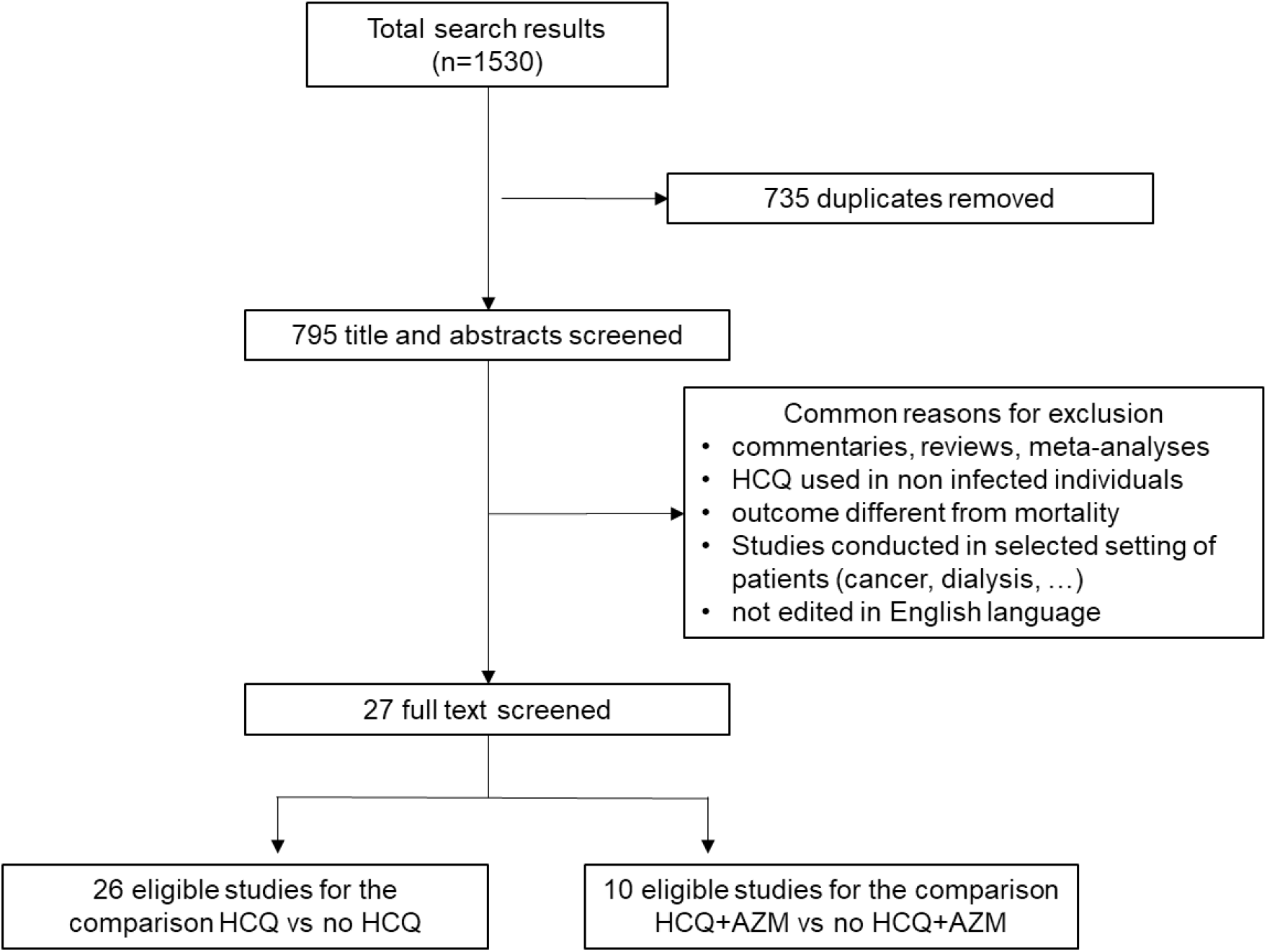
Flow diagram for study selection. HCQ means hydroxychloroquine; AZM means azithromycin

To be included in this meta-analysis, the study had to meet the following criteria: (1) clinical trials or cross-sectional studies or cohort studies; (2) quantitatively investigating the difference in mortality risk in unselected COVID-19 patients according to use or not of HCQ.

Twenty-seven articles were identified [7,9,11,12,14-36]. For ten of them [11,15,17,19,21,23,25,31-33] it was possible to extract data necessary for comparing HCQ+AZM *versus* no HCQ+AZM. For the other studies, it was not possible to systematically distinguish if HCQ therapy was complemented or not with AZM. The RCT by Mitjà et al. [29] was not included since the Authors found zero deaths both in HCQ and control group. The study of Mehra et al. [8] was excluded since the Authors retracted it owing to several concerns on the veracity of the data [10].

### Data Extraction and data analysis

For each study, odds ratio (OR) or hazard ratio (HR) and/or number of events (number of deaths and number of total COVID-19 patients) in both the HCQ (or HCQ+AZM) and respective control groups were extracted. If available, measure of association adjusted for covariates were retrieved. Number of events were used to calculate relative risk and 95% confidence intervals (CIs) when other measures of association were not available from the primary study. The following information was also extracted: study design, if the article was not peer-reviewed, region, level of adjustment, sample size, mortality rate in the entire cohort, percentage of patients treated with HCQ, mean duration of the treatment, mean daily dose after the first day and mean total dose of HCQ used. The total dose of HCQ was calculated as the sum of the amount of drug used in the first day plus daily dose multiplied by number of days of treatment after the first.

Pre-specified subgroup analyses have been conducted for all the additional characteristics retrieved. All analyses were performed using standard statistical procedures provided in RevMan5.1 (The Cochrane Collaboration, Oxford, United Kingdom). Data were combined using the general variance-based method, that requires information on the relative risk (or OR or HR) estimate and their 95% CI for each study. 95% CI were used to assess the variance and the relative weight of each study. Heterogeneity was assessed using the Higgin’s I^2^ metric. Fixed and random effects were considered, but due to the large heterogeneity observed, findings from random effects were considered as primary analysis. The hypothesis that publication bias might have affected the validity of the estimates was visually tested by a funnel plot–based approach.

## RESULTS

### Characteristics of the studies

The workflow of the process of study selection is reported in Figure 1. A total of 27 articles were found in the search. Twenty-six of these eligible studies were enrolled for analysing the association with mortality of HCQ use in patients with COVID-19, and 10 of them were valuable for analysing the association of HCQ+AZM.

The main characteristics of the 26 studies included in the meta-analysis are shown in Table 1. We found 4 RCT studies [9,14,17,34] and 22 observational studies; 5 articles were not published in peer reviewed journals; 3 studies reported not adjusted measure of association between HCQ and mortality; 13 studies have been conducted in Europe, 9 in North America and 3 in other countries. The outcome considered was the mortality for any cause; for the large majority of the studies, the mortality was intra-hospital. In all studies the control group was formed by patients without HCQ exposure (HCQ or HCQ+AZM). All studies included adult men and women COVID-19 patients, with the exception of two RCT [10,25], that included a portion of individuals with uncertain positivity to Sars-CoV-2. A total of N=44,521 COVID-19 patients (including N=7,324 from the 4 RCTs) were counted in the meta-analysis of HCQ, and N=12,153 in the meta-analysis of HCQ+AZM.

**Table 1.**
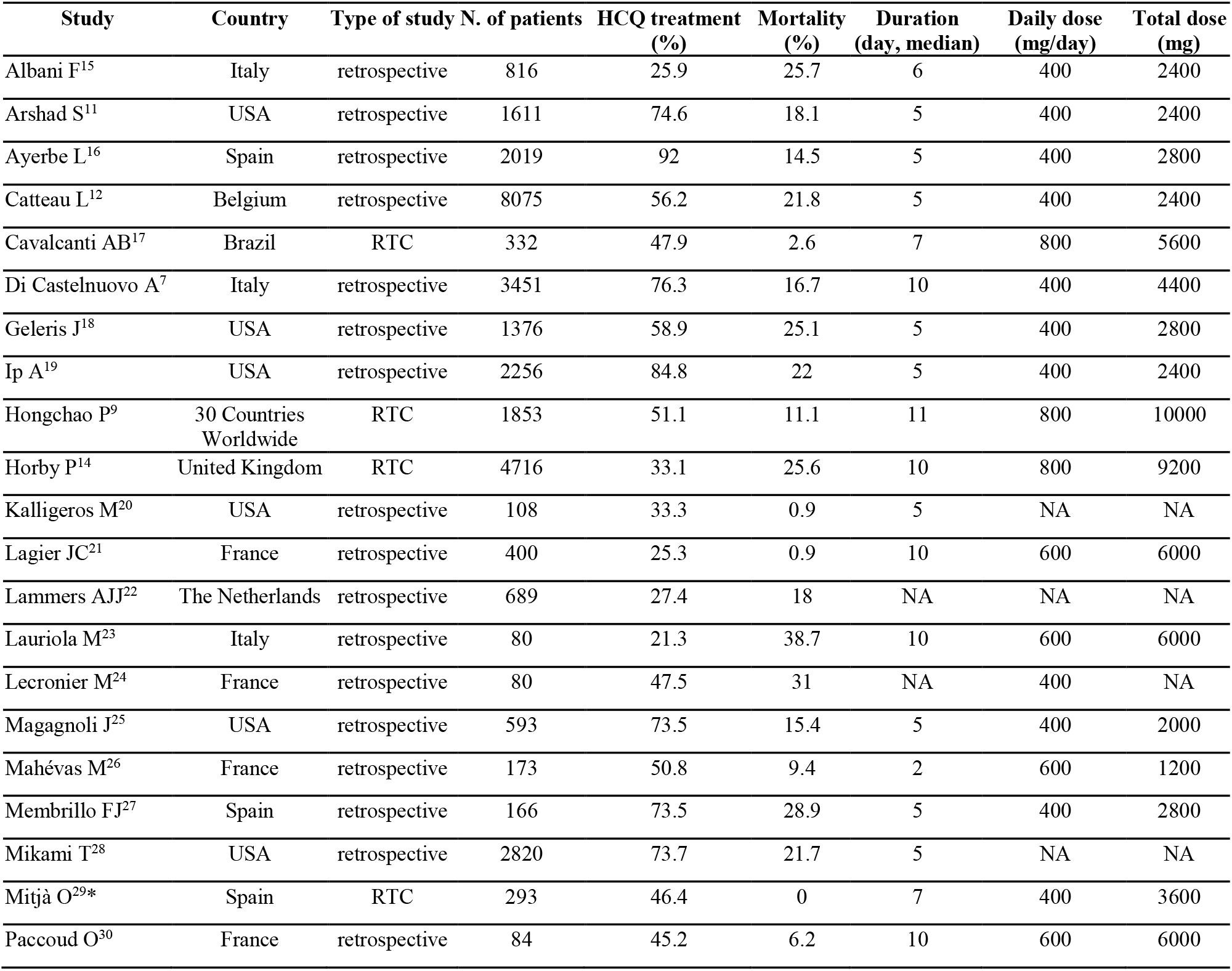
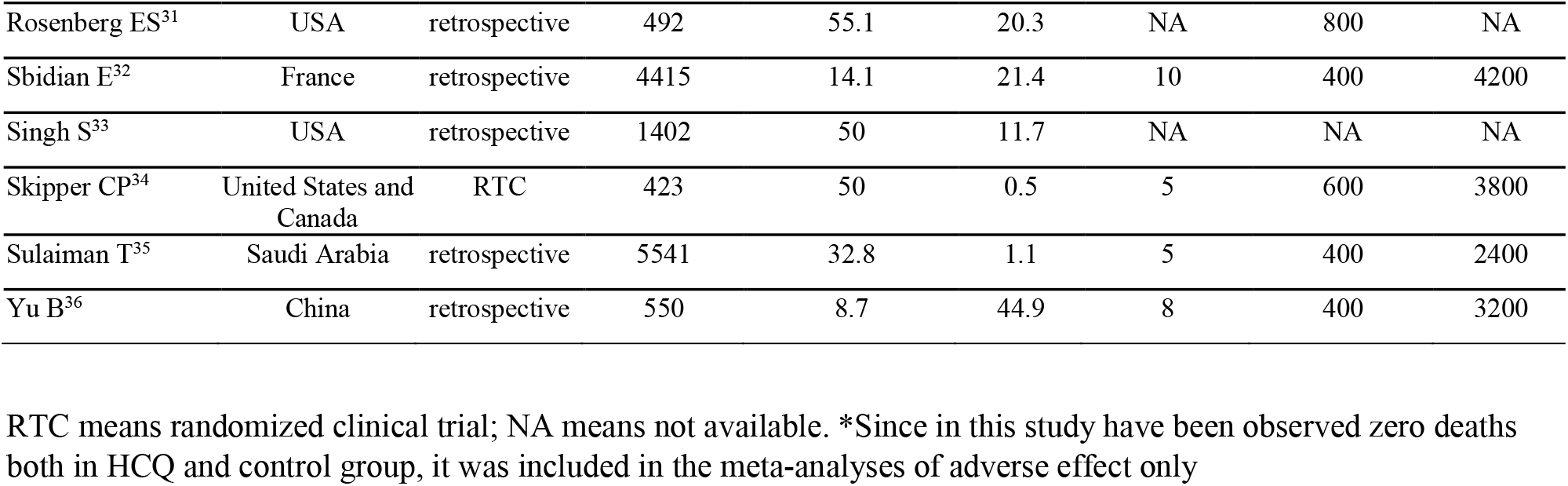
Characteristics of the studies included in the meta-analysis

| Study | Country | Type of study | N. of patients | HCQ treatment (%) | Mortality (%) | Duration (day, median) | Daily dose (mg/day) | Total dose (mg) |
| --- | --- | --- | --- | --- | --- | --- | --- | --- |
| Albani F<sup>15</sup> | Italy | retrospective | 816 | 25.9 | 25.7 | 6 | 400 | 2400 |
| Arshad S<sup>11</sup> | USA | retrospective | 1611 | 74.6 | 18.1 | 5 | 400 | 2400 |
| Ayerbe L<sup>16</sup> | Spain | retrospective | 2019 | 92 | 14.5 | 5 | 400 | 2800 |
| Catteau L<sup>12</sup> | Belgium | retrospective | 8075 | 56.2 | 21.8 | 5 | 400 | 2400 |
| Cavalcanti AB<sup>17</sup> | Brazil | RTC | 332 | 47.9 | 2.6 | 7 | 800 | 5600 |
| Di Castelnuovo A<sup>7</sup> | Italy | retrospective | 3451 | 76.3 | 16.7 | 10 | 400 | 4400 |
| Geleris J<sup>18</sup> | USA | retrospective | 1376 | 58.9 | 25.1 | 5 | 400 | 2800 |
| Ip A<sup>19</sup> | USA | retrospective | 2256 | 84.8 | 22 | 5 | 400 | 2400 |
| Hongchao P<sup>9</sup> | 30 Countries Worldwide | RTC | 1853 | 51.1 | 11.1 | 11 | 800 | 10000 |
| Horby P<sup>14</sup> | United Kingdom | RTC | 4716 | 33.1 | 25.6 | 10 | 800 | 9200 |
| Kalligeros M<sup>20</sup> | USA | retrospective | 108 | 33.3 | 0.9 | 5 | NA | NA |
| Lagier JC<sup>21</sup> | France | retrospective | 400 | 25.3 | 0.9 | 10 | 600 | 6000 |
| Lammers AJJ<sup>22</sup> | The Netherlands | retrospective | 689 | 27.4 | 18 | NA | NA | NA |
| Lauriola M<sup>23</sup> | Italy | retrospective | 80 | 21.3 | 38.7 | 10 | 600 | 6000 |
| Lecronier M<sup>24</sup> | France | retrospective | 80 | 47.5 | 31 | NA | 400 | NA |
| Magagnoli J<sup>25</sup> | USA | retrospective | 593 | 73.5 | 15.4 | 5 | 400 | 2000 |
| Mahévas M<sup>26</sup> | France | retrospective | 173 | 50.8 | 9.4 | 2 | 600 | 1200 |
| Membrillo FJ<sup>27</sup> | Spain | retrospective | 166 | 73.5 | 28.9 | 5 | 400 | 2800 |
| Mikami T<sup>28</sup> | USA | retrospective | 2820 | 73.7 | 21.7 | 5 | NA | NA |
| Mitjà O<sup>29*</sup> | Spain | RTC | 293 | 46.4 | 0 | 7 | 400 | 3600 |
| Paccoud O<sup>30</sup> | France | retrospective | 84 | 45.2 | 6.2 | 10 | 600 | 6000 |
| Rosenberg ES<sup>31</sup> | USA | retrospective | 492 | 55.1 | 20.3 | NA | 800 | NA |
| Sbidian E<sup>32</sup> | France | retrospective | 4415 | 14.1 | 21.4 | 10 | 400 | 4200 |
| Singh S<sup>33</sup> | USA | retrospective | 1402 | 50 | 11.7 | NA | NA | NA |
| Skipper CP<sup>34</sup> | United States and<br>Canada | RTC | 423 | 50 | 0.5 | 5 | 600 | 3800 |
| Sulaiman T<sup>35</sup> | Saudi Arabia | retrospective | 5541 | 32.8 | 1.1 | 5 | 400 | 2400 |
| Yu B<sup>36</sup> | China | retrospective | 550 | 8.7 | 44.9 | 8 | 400 | 3200 |

RTC means randomized clinical trial; NA means not available. \*Since in this study have been observed zero deaths both in HCQ and control group, it was included in the meta-analyses of adverse effect only

### Pooled analysis, HCQ as exposure

Forest plot on the association between HCQ and mortality is reported in Figure 2. Pooling of data from 22 observational studies, which accounted for 86.2% of the total weight, the use of HCQ has been associated with 25% lower mortality risk (pooled risk ratio: 0.75, 95%CI: 0.63 to 0.89; high level of heterogeneity: I^2^=80%, random effects). Conversely, the association was not evident in the 4 RCTs (13.8% of the weight; pooled risk ratio: 1.11, 95%CI: 0.99 to 1.24; I^2^=0%). Grand total by pooling of both RCTs and observational studies (weight 100%) pointed to a relative reduction of mortality in support of HCQ by 7% to 33% (figure 2).

**Figure 2.**
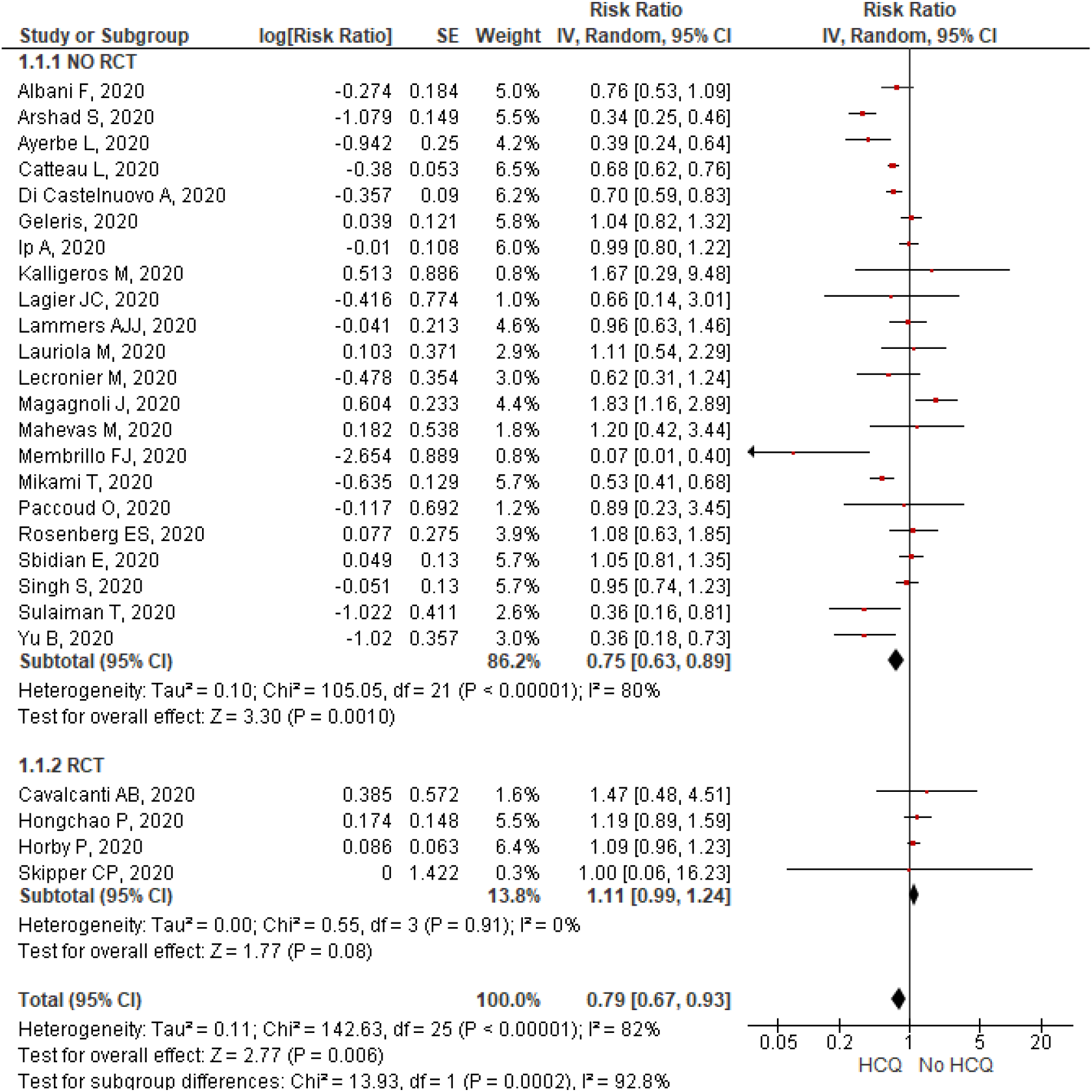
Forest plot for association of hydroxychloroquine use with COVID-19 mortality (random effects) RCT means randomized clinical trial; SE means standard error; HCQ means hydroxychloroquine

Subgroup analyses according to main features of primary studies are presented in Table 2. The association of HCQ with lower mortality was observed with very low differences in all subgroups, with the exception of dose grouping.

**Table 2.**
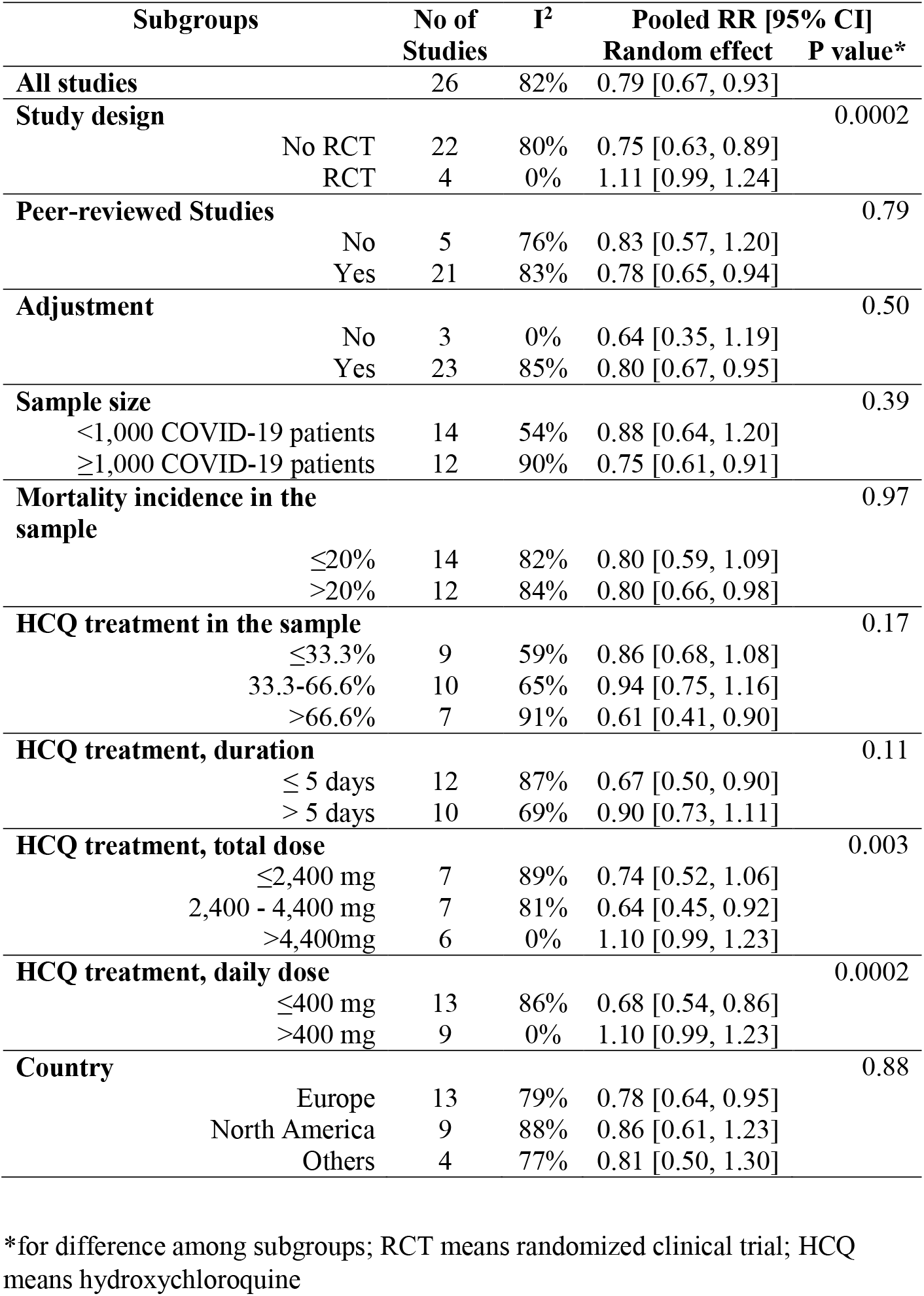
Pooled analysis in different subgroups of studies

The reduced mortality was in fact confined to studies that used a daily dose ≤400 mg (as estimated in days of treatment after the first, in which a higher (double for most) dose of drug was administered); pooling n=9 studies which used more than 400 mg of HCQ daily resulted in an overall measure of association equal to 1.10 (95%CI: 0.99 to 1.23; table 2 and supplementary figure 1). Also, the magnitude of the association was higher in studies which used HCQ for 5 or less days (table 2 and supplementary figure 2). Pooling studies which used more than 4,400 mg of HCQ during the entire phase of treatment set an overall measure of association equal to 1.10 (95%CI: 0.99 to 1.23; table 2 and supplementary figure 3). These findings were confirmed in subgroup analyses restricted to observational studies: the pooled measure of association was equal to 0.98 (95%CI: 0.54 to 1.77) and 1.05 (95%CI: 0.73 to 1.53) in studies with a total HCQ dose >4,400 mg (n=3) or a daily HCQ dose >400 mg (n=5), respectively.

### Pooled analysis, HCQ+AZM as exposure

Figure 3 reported random forest for 10 studies comparing HCQ+AZM. Use of the combination HCQ+AZM was associated with 25% lower mortality risk, with very large uncertainty (pooled risk ratio: 0.75, 95%CI: 0.50 to 1.13; P for testing of overall effect=0.17; high level of heterogeneity: I^2^=91%, random effects).

**Figure 3.**
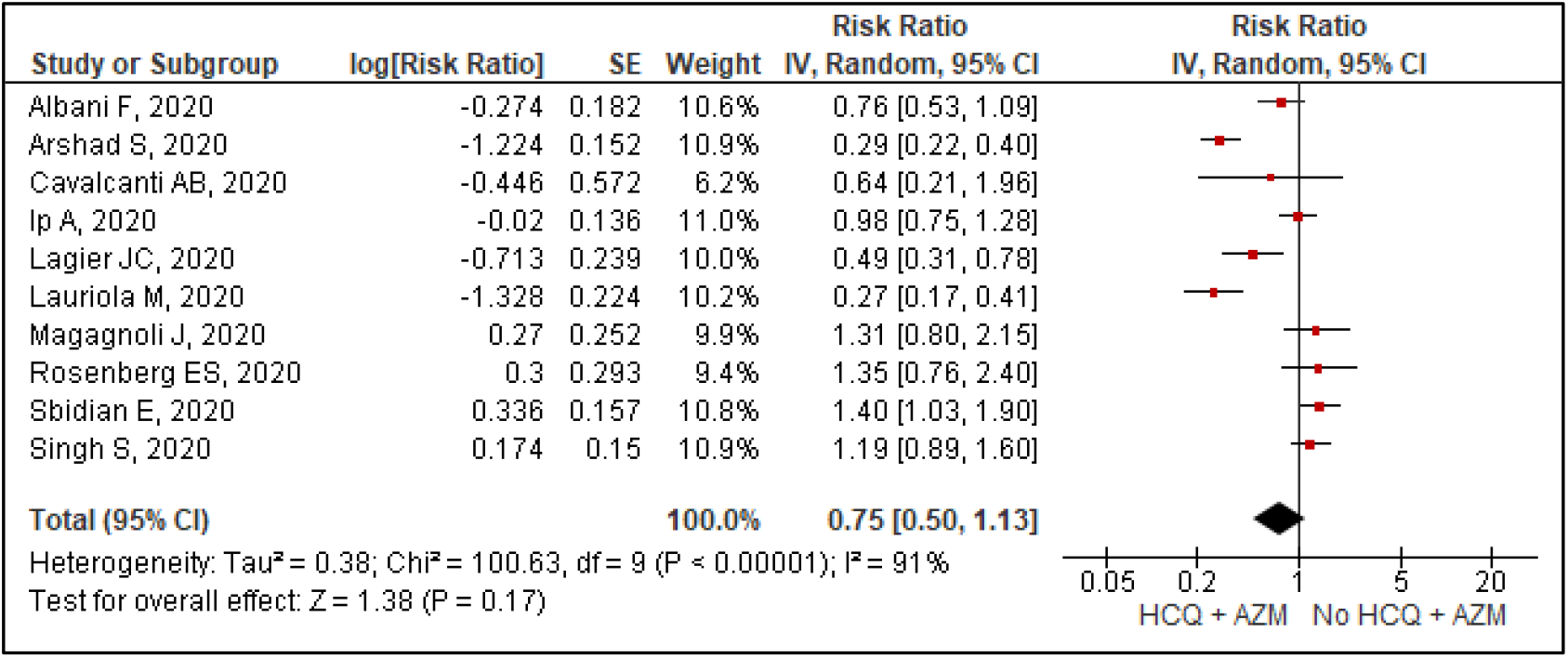
Forest plot for association of hydroxychloroquine + azithromycin use with COVID-19 mortality (random effects) HCQ means hydroxychloroquine; AZM means azithromycin

By visual inspection of funnel plots (Supplementary Figure 4), we failed to observe any selection bias for both meta-analyses.

### Pooled analysis of adverse events

Pooled analyses of RCTs on the relationship between HCQ and incidence of adverse effects are reported in supplementary figure 5. HCQ use was associated with an increased risk of adverse effects of any type (panel A). On the contrary, patients treated with HCQ in RCTs showed a similar rate of serious adverse events (panel B), as that non-treated with HCQ (pooled risk ratio: 1.16, 95%CI: 0.86 to 1.55; P for testing of overall effect=0.33).

## DISCUSSION

In a meta-analysis of 26 studies (4 RCTs) involving 44,814 COVID-19 patients, the use of HCQ was associated with a 21% lower risk of total mortality. The association was apparent by pooling 22 observational studies and was more evident in studies which used lower HCQ doses. No association was found pooling 4 RCTs. Use of HCQ was not associated with severe adverse events.

The potential for selection bias in observational studies is not negligible. The decision from the clinician to utilize or not a drug may depend on comorbidities and baseline risk of the patient. However, in the pandemic, and in absence of guidelines and specific anti COVID-19 drugs, allocation of HCQ in observational studies was not associated systematically with a lower or higher baseline risk profile. For example, in the CORIST study [6] patients receiving HCQ were more likely younger and less likely had ischemic heart disease, cancer or chronic kidney disease, but, on the contrary, they were more likely men and had higher levels of C-reactive protein. As a consequence, it is not clear if in that particular study HCQ patients were potentially at higher or lower risk of a negative prognosis. In attempting to account for baseline differences between patients who received HCQ and those who did not, we used the results for adjusted measure of association for each study, and this was possible for 22 out of 26 studies included in the meta-analysis. After the exclusion of 3 unadjusted studies [16,21,24], the strength of the overall association of HCQ with mortality was merely reduced from 0.79 to 0.80. Although we attempted to control for potential confounding factors inherent to patient and clinical characteristics, it is possible that unmeasurable confounding still remains, and this may explain the different finding between observational and RCT studies. However, it is hard to determine which are, if any, the unmeasured characteristics that have confused so strongly the association between HCQ and mortality in COVID-19 that was observed in observational studies. In fact, these features must be a) unmeasured in observational studies; b) associated with mortality in COVID-19 and c) associated with HCQ use, in a way that when the risky conditions are present the clinicians tend systematically to avoid using HCQ. For example, HCQ is contraindicated in patients with cardiomyopathy but this condition has been mostly measured in observational studies and was not recognised as a risk factor for mortality in COVID-19 patients.

The dissimilar findings between observational and RCTs we found might also be explained by differences in HCQ dosage [37]. Interestingly, we observed that the reduced mortality associated with HCQ treatment is actually confined to studies that used a daily dose ≤400 mg, or a total dose ≤4,400 mg or which used HCQ for 5 or less days. Obviously these three conditions largely overlapped in studies, that we can now designate as “at low HCQ dose”. Remarkably, none of the 4 RCTs are in this category. In detail, the RECOVERY [14] and the SOLIDARITY study [9] used 800 mg/day for 9 or 10 days (after the first), respectively and a total dose of 9200 or 10000 mg of HCQ (including the dose at first day) respectively, a very high dose regimen as confronted to the rest of studies, particularly of the observational ones.

The possibility that HCQ reduced the risk of negative prognosis in COVID-19 patients when only administered at “low dose” cannot be here undoubtedly proven starting from our findings, but it is a plausible hypothesis that may explain the different result between observational and RCT studies and, more importantly, might be useful in disentangling the debate on HCQ use in COVID-19.

High levels of HCQ administration were used in RCTs to maximise the antiviral activity of the drug that was considered to be the main mechanism of action of HCQ in this context. In some studies, the inverse association of HCQ with inpatient mortality was more evident in elderly, in patients who experienced a higher degree of COVID-19 severity or having elevated C-reactive protein levels [7], suggesting that the anti-inflammatory potential of HCQ may have had a more important role than its antiviral properties. HCQ, indeed, beside an antiviral activity, may have both anti-inflammatory and anti-thrombotic effects [3]. This can justify its effect in reducing mortality risk, since Sars-Cov-2 can induce pulmonary microthrombi and coagulopathy, that are a possible cause of its severity [38, 39] and the lack in preventing SARS-CoV-2 infection after exposure [40]. On the other end, National guidelines suggested to use HCQ 200 mg twice daily for 5-7 days probably to maintain a better risk benefit profile hypothesizing that low doses could be more effective and safer. Indeed, non-sigmoidal, bell-shaped dose-response curves are possible with drugs having complex biological effects, multiple-binding sites or cellular and organ targets. On the other hand, anti SARS-2-CoV-2 activity of HCQ has been confirmed in Vero cells [41]. HCQ is also reported to reduce secretion of IFN-γ and IL-17 in activated Th1 and Th17 cells, respectively [42].

The concomitant use of azithromycin seems to not neither increase nor decrease the effect, if any, of the HCQ since the combination of the two drugs was associated with a lower mortality risk at very similar extent to that observed for HCQ alone, but the assumption is inconclusive because of the very large uncertainty in the findings.

A main concern with HCQ treatment have been its side effects, in particular a severe cardiovascular toxicity. Indeed, HCQ can cause prolongation of the QT interval on electrocardiogram [43], which could be exacerbated by coadministration with azithromycin, widely prescribed as co-treatment in Covid-19 treatment. Our meta-analysis of data from RCTs, that allowed a proper evaluation of side effects, shows that use of HCQ was associated with an increase in side effects of any type, but not of major type, including cardiovascular events. This despite the high prevalence of cardiovascular disease in patients with COVID-19 or the high dose used in RTCs.

This meta-analysis has the strength of including all available data recently published and that have not been included in previous meta-analyses [13,44], and of considering modification of effect by dosing of HCQ. As major limitations, we recognised that majority of the primary studies were observational, the pooled findings suffer of a high degree of heterogeneity and that results in observational and RCT studies were different.

### Conclusion

In conclusion, HCQ was not associated with increased or decreased mortality in COVID-19 patients when only RCTs were pooled, but it was associated with 8% to 35% reduced mortality when observational studies were also included. The association was mainly apparent by pooling studies using lower doses of HCQ. Use of HCQ was not associated with severe adverse events.

These results should be considered with caution, because the majority of the studies included were observational and retrospective and the possibility of confounding could not be fully excluded. However, at present, this is the largest comprehensive quantitative overview on the association of HCQ with mortality in COVID-19 patients, and our findings underscoring HCQ dosage effects can help disentangling the debate on HCQ use and encourage the planning of RCTs using low doses of HCQ in hospitalised COVID-19 patients.

## Data Availability

The manuscript is a meta-analysis based on available data

## Acknowledgments

SC was the recipient of a Fondazione Umberto Veronesi Travel Grant.

## Author contributions

ADC and LI contributed to the conception and design of the work and interpretation of data; SC and ADC managed study selection and data extraction and critically reviewed the results; SC analysed the data; ADC and LI wrote the paper; LI, AC, RC and GdG originally inspired the research and critically reviewed the manuscript.

All Authors approved the final version of the manuscript.

---

**Supplementary Figure 1.**
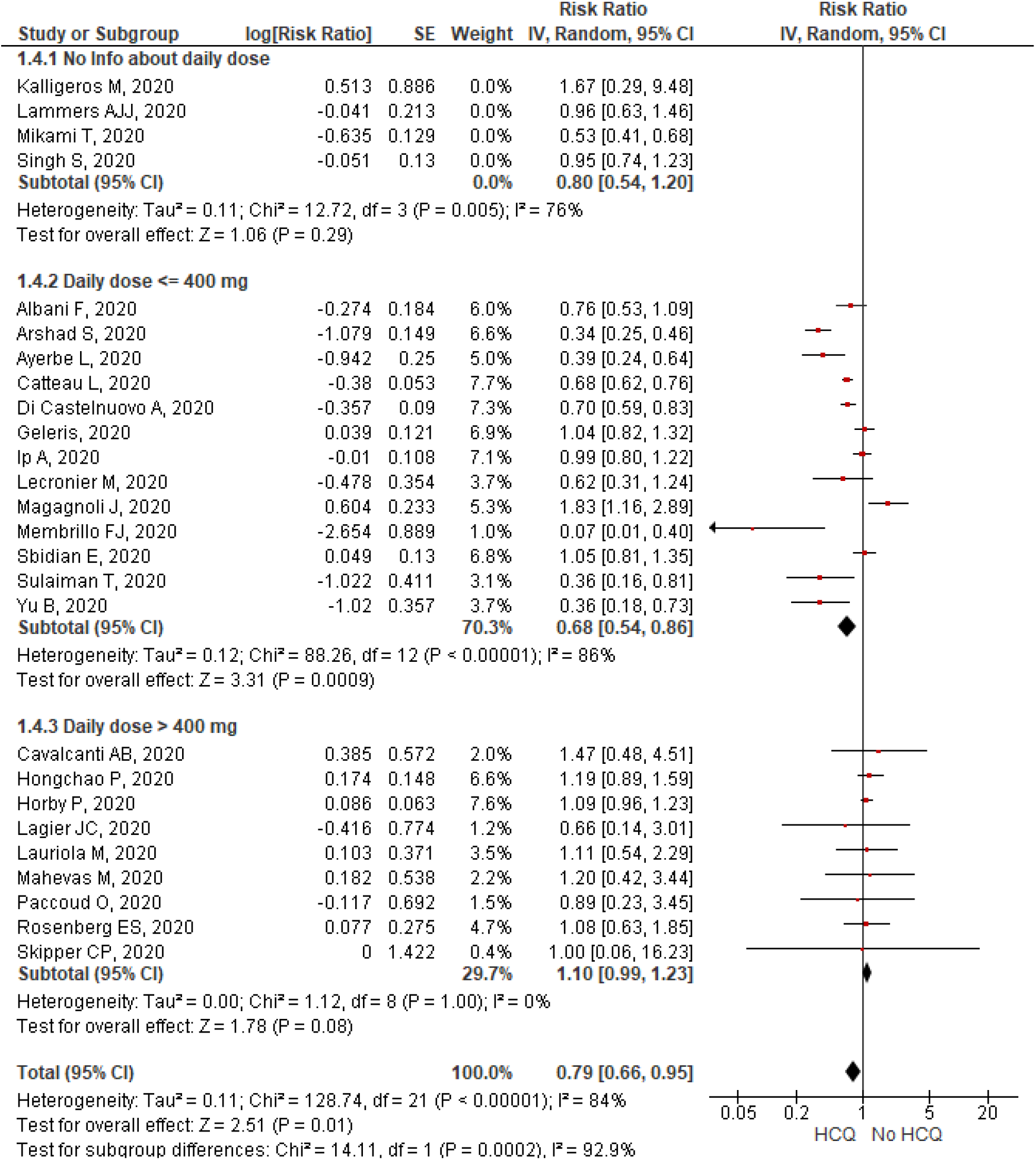
Forest plot for association of hydroxychloroquine use with COVID-19 mortality (random effects), according to daily dose (as estimated in the days of treatment following the first)

**Supplementary Figure 2.**
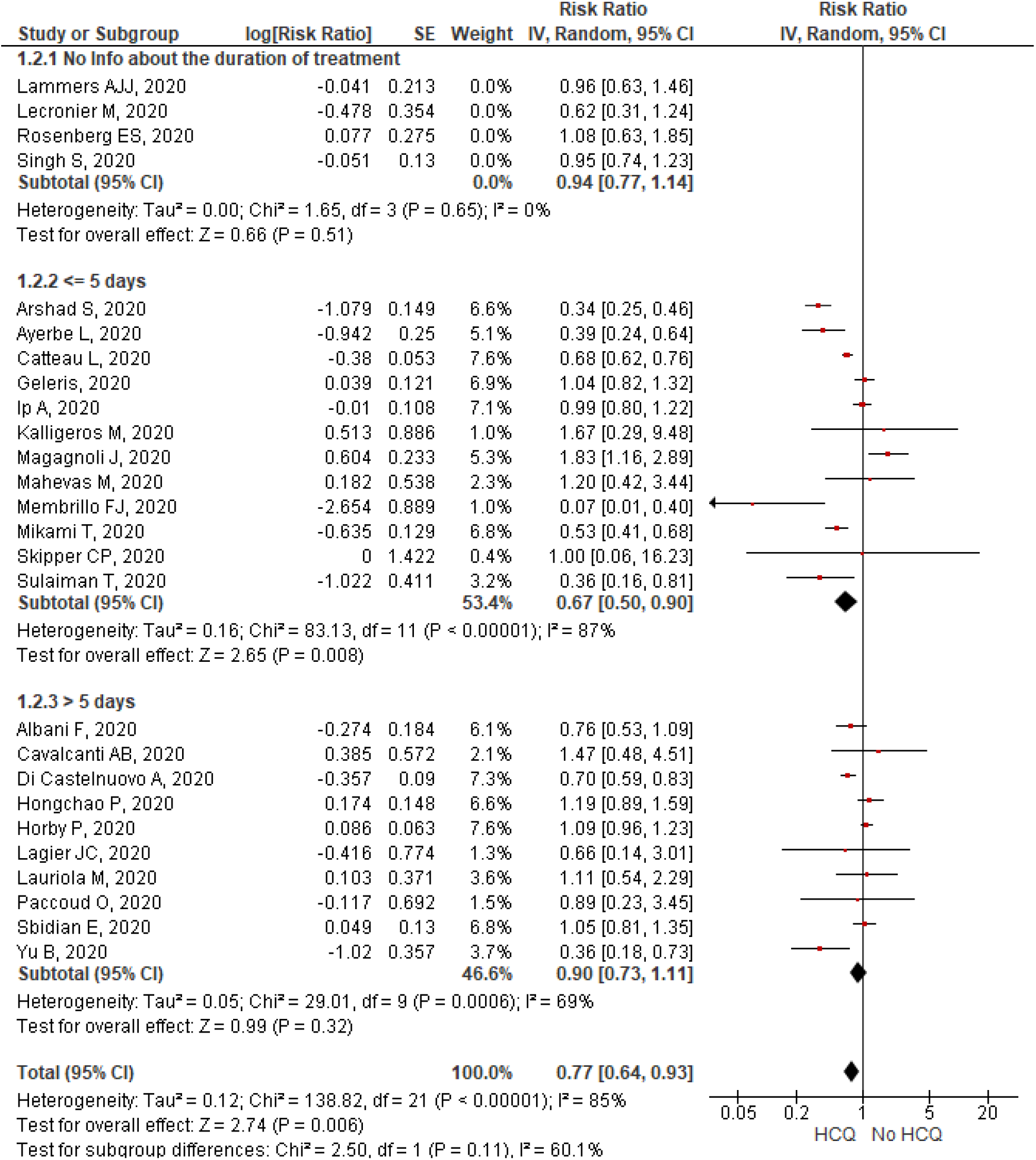
Forest plot for association of hydroxychloroquine use with COVID-19 mortality (random effects), according to duration of treatment.

**Supplementary Figure 3.**
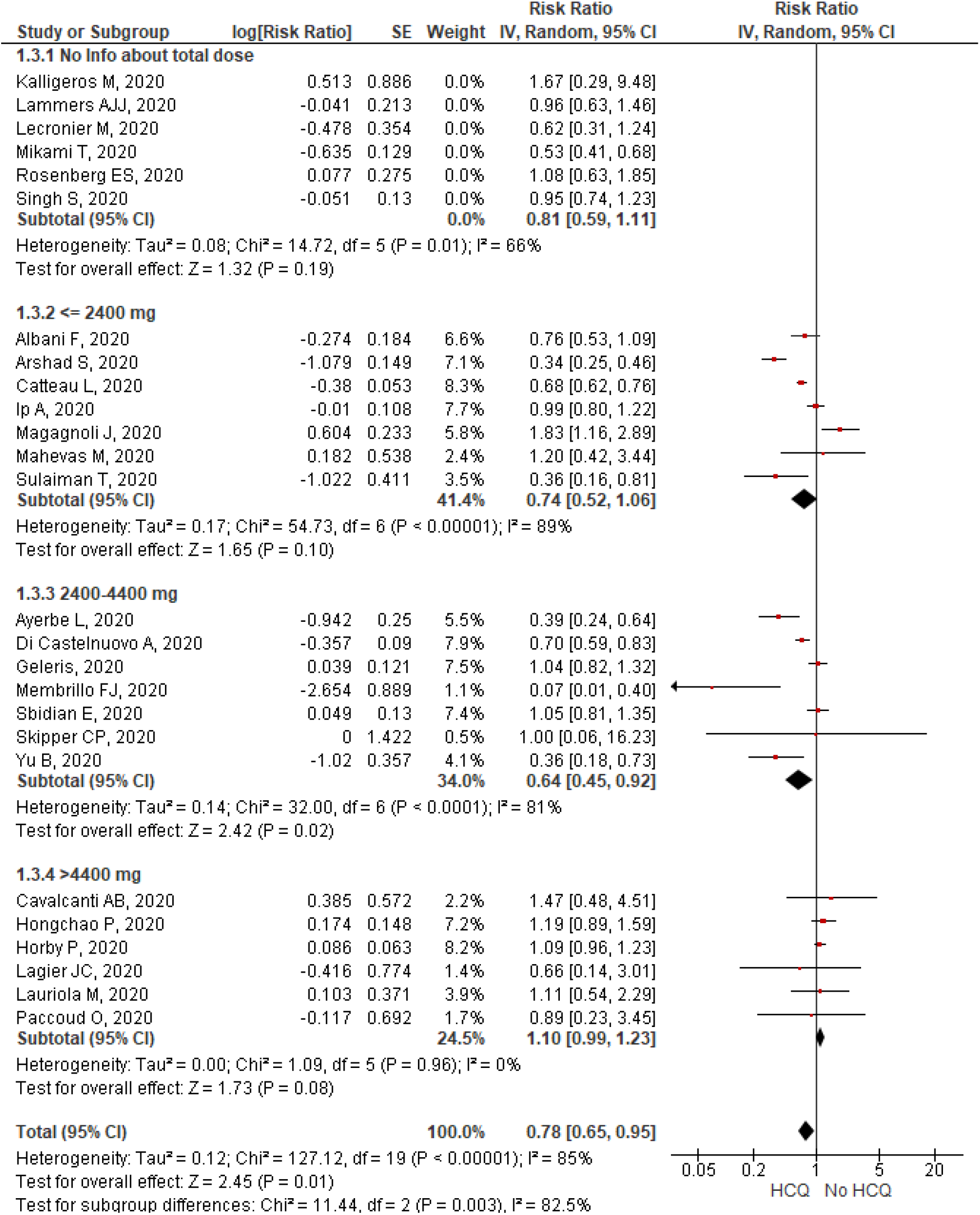
Forest plot for association of hydroxychloroquine use with COVID-19 mortality (random effects), according to total dose (calculated as the sum of the amount of drug used in the first day plus daily dose multiplied by number of days of treatment following the first)

**Supplementary Figure 4.**
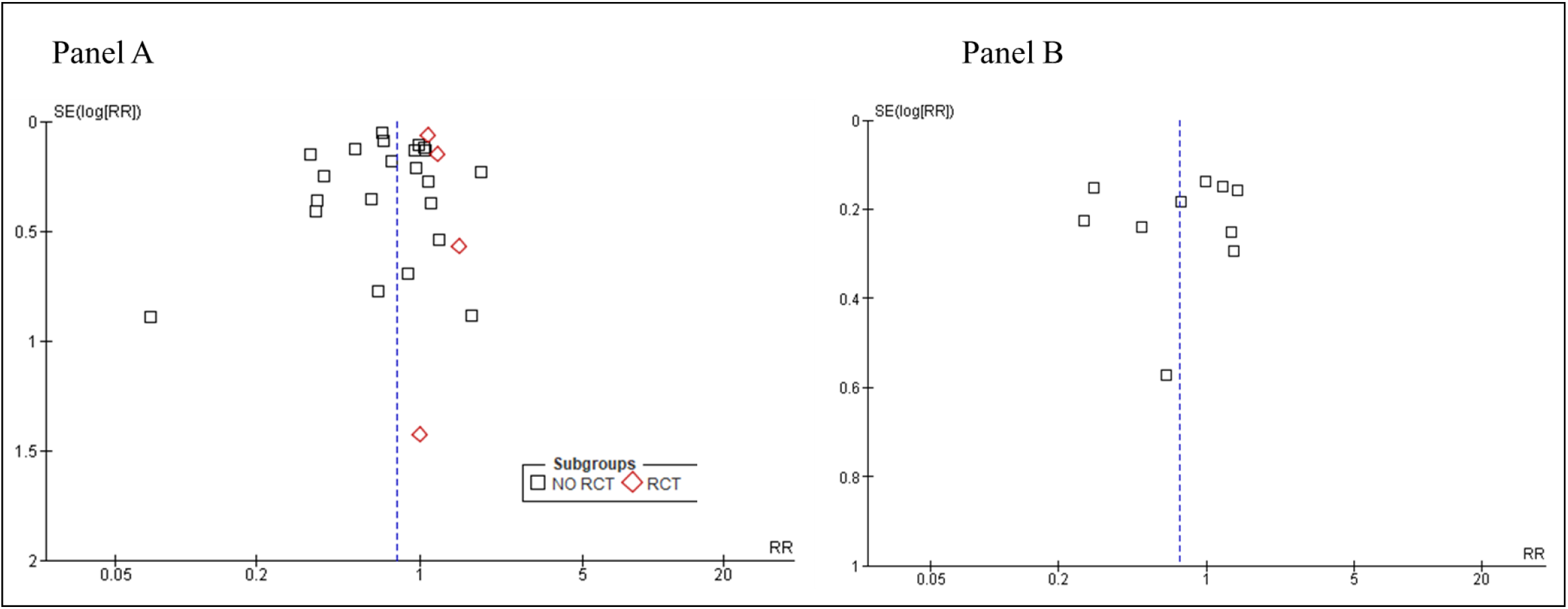
Forest plot for association between HCQ treatment (panel A) or HCQ+AZM (panel B) and mortality in COVID-19 patients. RCT mean randomized controlled trial; RR means relative risk; SE means standard error; HCQ means hydroxychloroquine; AZM means azithromycin

**Supplementary Figure 5.**
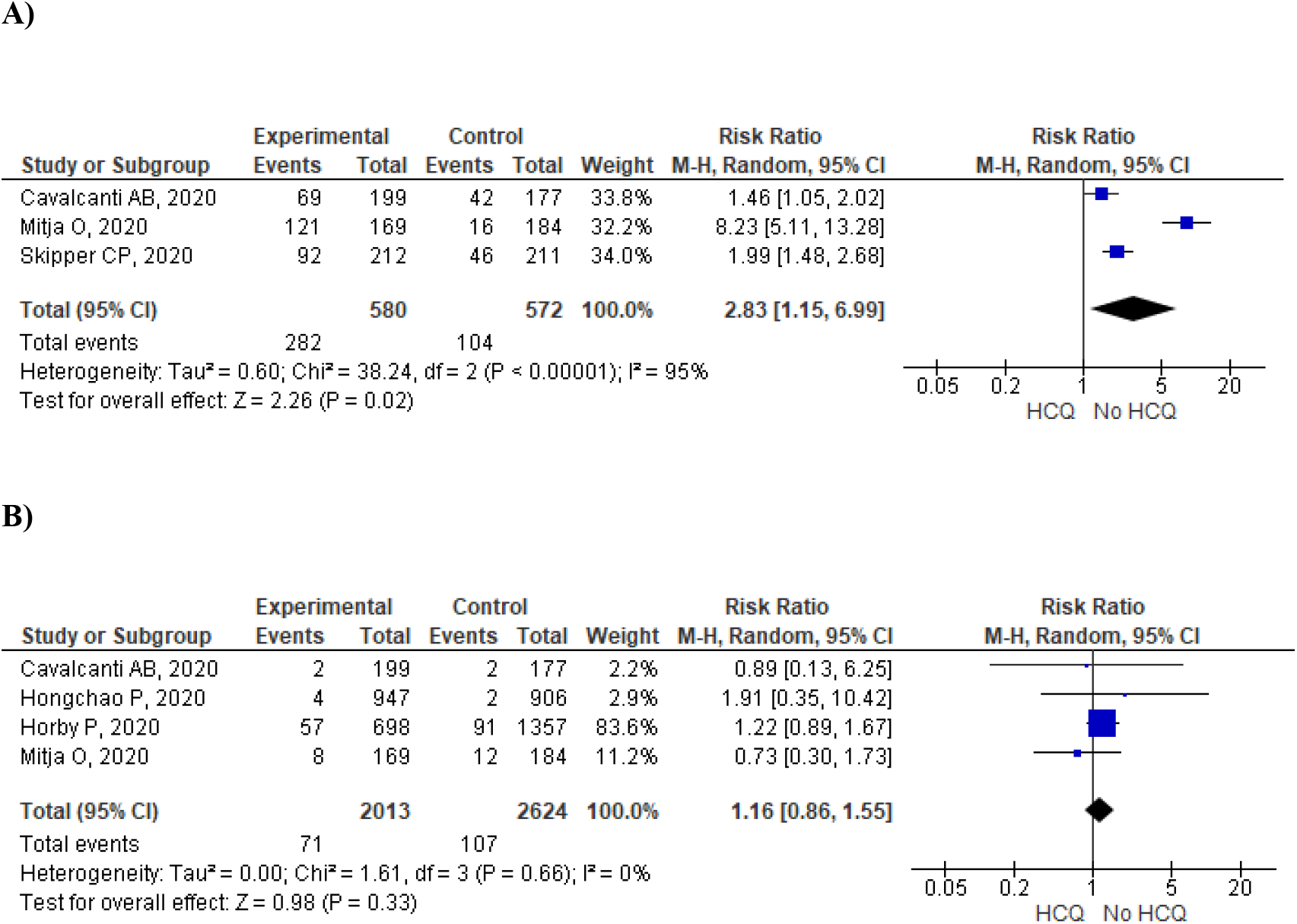
Forest plot for association of hydroxychloroquine use with any (panel A) or major (panel B) adverse effects. Data are from randomised clinical trials. The study by Mitja et al. (ref=meta esclusi 3) was not included in the meta-analysis on mortality since the authors found no deaths both in HCQ and control group.

## REFERENCES

1) Shukla AM, Wagle Shukla A. Expanding horizons for clinical applications of chloroquine, hydroxychloroquine, and related structural analogues. Drugs Context. 2019;8:2019-9-1. Published 2019 Nov 25. doi:10.7573/dic.2019-9-1

2) Savarino A, Boelaert JR, Cassone A, Majori G, Cauda R. Effects of chloroquine on viral infections: an old drug against today’s diseases?. Lancet Infect Dis. 2003;3(11):722–727. doi:10.1016/s1473-3099(03)00806-5

3) Quiros Roldan E, Biasiotto G, Magro P, Zanella I. The possible mechanisms of action of 4- aminoquinolines (chloroquine/hydroxychloroquine) against Sars-Cov-2 infection (COVID- 19): A role for iron homeostasis? [published online ahead of print, 2020 May 13]. Pharmacol Res. 2020;158:104904. doi:10.1016/j.phrs.2020.104904

4) Dowall SD, Bosworth A, Watson R, et al. Chloroquine inhibited Ebola virus replication in vitro but failed to protect against infection and disease in the in vivo guinea pig model. J Gen Virol. 2015;96(12):3484–3492. doi:10.1099/jgv.0.000309

5) Savarino A, Shytaj IL. Chloroquine and beyond: exploring anti-rheumatic drugs to reduce immune hyperactivation in HIV/AIDS. Retrovirology. 2015;12:51. Published 2015 Jun 18. doi:10.1186/s12977-015-0178-0

6) de Wilde AH, Jochmans D, Posthuma CC, et al. Screening of an FDA-approved compound library identifies four small-molecule inhibitors of Middle East respiratory syndrome coronavirus replication in cell culture. Antimicrob Agents Chemother. 2014;58(8):4875–4884. doi:10.1128/AAC.03011-14

7) COVID-19 RISK and Treatments (CORIST) Collaboration. Di Castelnuovo A, Costanzo S, Antinori A, et al. Use of hydroxychloroquine in hospitalised COVID-19 patients is associated with reduced mortality: Findings from the observational multicentre Italian CORIST study.Eur J Intern Med. 2020 Aug 25:S0953-6205(20)30335-6. doi: 10.1016/j.ejim.2020.08.019. Epub ahead of print. PMID: 32859477; PMCID: PMC7446618.

8) Mehra MR, Desai SS, Ruschitzka F, Patel AN. Hydroxychloroquine or chloroquine with or without a macrolide for treatment of COVID-19: a multinational registry analysis. Lancet. 2020 May 22. Retracted article

9) WHO Solidarity Trial Consortium, Hongchao P, Richard P, Quarraisha AK, et al. Repurposed antiviral drugs for COVID-19; interim WHO SOLIDARITY trial results. https://doi.org/10.1101/2020.10.15.20209817.

10) Mehra MR, Ruschitzka F, Patel AN. Retraction—Hydroxychloroquine or chloroquine with or without a macrolide for treatment of COVID-19: a multinational registry analysis. Lancet. 2020 June 5. DOI:https://doi.org/10.1016/S0140-6736(20)31324-6

11) Arshad S, Kilgore P, Chaudhry ZS, et al. Treatment with hydroxychloroquine, azithromycin, and combination in patients hospitalized with COVID-19. Int J Infect Dis. 2020 Aug;97:396–403. doi: 10.1016/j.ijid.2020.06.099. Epub 2020 Jul 2. PMID: 32623082; PMCID: PMC7330574.

12) Catteau L, Dauby N, Montourcy M, et al. Low-dose hydroxychloroquine therapy and mortality in hospitalised patients with COVID-19: a nationwide observational study of 8075 participants. Int J Antimicrob Agents. 2020 Oct;56(4):106144. doi: 10.1016/j.ijantimicag.2020.106144. Epub 2020 Aug 24. PMID: 32853673; PMCID: PMC7444610.

13) Fiolet T, Guihur A, Rebeaud ME, Mulot M, Peiffer-Smadja N, Mahamat-Saleh Y. Effect of hydroxychloroquine with or without azithromycin on the mortality of coronavirus disease 2019 (COVID-19) patients: a systematic review and meta-analysis. Clin Microbiol Infect. 2020 Aug 26:S1198-743X(20)30505-X. doi: 10.1016/j.cmi.2020.08.022. Epub ahead of print. PMID: 32860962; PMCID: PMC7449662.

14) RECOVERY Collaborative Group. Horby P, Mafham M, Linsell L, et al. Effect of Hydroxychloroquine in Hospitalized Patients with Covid-19. N Engl J Med. 2020 Oct 8. doi: 10.1056/NEJMoa2022926. Epub ahead of print. PMID: 33031652.

15) Albani F, Fusina F, Giovannini A, et al. Impact of Azithromycin and/or Hydroxychloroquine on Hospital Mortality in COVID-19. J Clin Med. 2020 Aug 30;9(9):E2800. doi: 10.3390/jcm9092800. PMID: 32872629.

16) Ayerbe L, Risco-Risco C, Ayis S. The association of treatment with hydroxychloroquine and hospital mortality in COVID-19 patients. Intern Emerg Med. 2020 Sep 30. doi: 10.1007/s11739-020-02505-x. Epub ahead of print. PMID: 32997237.

17) Cavalcanti AB, Zampieri FG, Rosa RG, et al. Hydroxychloroquine with or without Azithromycin in Mild-to-Moderate Covid-19. N Engl J Med. 2020 Jul 23:NEJMoa2019014. doi: 10.1056/NEJMoa2019014. Epub ahead of print. PMID: 32706953; PMCID: PMC7397242.

18) Geleris J, Sun Y, Platt J, et al. Observational Study of Hydroxychloroquine in Hospitalized Patients with Covid-19. N Engl J Med. 2020 Jun 18;382(25):2411–2418. doi: 10.1056/NEJMoa2012410. Epub 2020 May 7. PMID: 32379955; PMCID: PMC7224609.

19) Ip A, Berry DA, Hansen E, et al. Hydroxychloroquine and tocilizumab therapy in COVID- 19 patients-An observational study. PLoS One. 2020 Aug 13;15(8):e0237693. doi: 10.1371/journal.pone.0237693. PMID: 32790733; PMCID: PMC7425928.

20) Kalligeros M, Shehadeh F, Atalla E, et al. Hydroxychloroquine use in hospitalised patients with COVID-19: An observational matched cohort study. J Glob Antimicrob Resist. 2020 Sep;22:842–844. doi: 10.1016/j.jgar.2020.07.018. Epub 2020 Aug 5. PMID: 32763357; PMCID: PMC7403006.

21) Lagier JC, Million M, Gautret P, et al. Outcomes of 3,737 COVID-19 patients treated with hydroxychloroquine/azithromycin and other regimens in Marseille, France: A retrospective analysis. Travel Med Infect Dis. 2020 Jul-Aug;36:101791. doi: 10.1016/j.tmaid.2020.101791. Epub 2020 Jun 25. PMID: 32593867; PMCID: PMC7315163.

22) Lammers AJJ, Brohet RM, Theunissen REP, et al. Early Hydroxychloroquine but not Chloroquine use reduces ICU admission in COVID-19 patients, International Journal of Infectious Diseases (2020), doi: https://doi.org/10.1016/j.ijid.2020.09.1460

23) Lauriola M, Pani A, Ippoliti G, et al. Effect of combination therapy of hydroxychloroquine and azithromycin on mortality in COVID-19 patients. Clin Transl Sci. 2020 Sep 14. doi: 10.1111/cts.12860. Epub ahead of print. PMID: 32926573.

24) Lecronier M, Beurton A, Burrel S, et al. Comparison of hydroxychloroquine, lopinavir/ritonavir, and standard of care in critically ill patients with SARS-CoV-2 pneumonia: an opportunistic retrospective analysis. Crit Care. 2020 Jul 11;24(1):418. doi: 10.1186/s13054-020-03117-9. PMID: 32653015; PMCID: PMC7351645.

25) Magagnoli J, Narendran S, Pereira F, et al. Outcomes of Hydroxychloroquine Usage in United States Veterans Hospitalized with COVID-19. Med (N Y). 2020 Jun 5. doi: 10.1016/j.medj.2020.06.001. Epub ahead of print. PMID: 32838355; PMCID: PMC7274588.

26) Mahévas M, Tran VT, Roumier M, et al. Clinical efficacy of hydroxychloroquine in patients with covid-19 pneumonia who require oxygen: observational comparative study using routine care data. BMJ. 2020 May 14;369:m1844. doi: 10.1136/bmj.m1844. Erratum in: BMJ. 2020 Jun 18;369:m2328. PMID: 32409486; PMCID: PMC7221472.

27) Membrillo de Novales FJ, Ramírez-Olivencia G, Estébanez M, et al. et al. Early Hydroxychloroquine Is Associated with an Increase of Survival in COVID-19 Patients: An Observational Study. Preprints 2020, 2020050057 doi: 10.20944/preprints202005.0057.v1.

28) Mikami T, Miyashita H, Yamada T, et al. Risk Factors for Mortality in Patients with COVID-19 in New York City. J Gen Intern Med. 2020 Jun 30:1–10. doi: 10.1007/s11606-020-05983-z. Epub ahead of print. PMID: 32607928; PMCID: PMC7325642.

29) Mitjà O, Corbacho-Monné M, Ubals M, et al. Hydroxychloroquine for Early Treatment of Adults with Mild Covid-19: A Randomized-Controlled Trial. Clin Infect Dis. 2020 Jul 16:ciaa1009. doi: 10.1093/cid/ciaa1009. Epub ahead of print. PMID: 32674126; PMCID: PMC7454406.

30) Paccoud O, Tubach F, Baptiste A, et al. Compassionate use of hydroxychloroquine in clinical practice for patients with mild to severe Covid-19 in a French university hospital. Clin Infect Dis. 2020 Jun 18:ciaa791. doi: 10.1093/cid/ciaa791. Epub ahead of print. PMID: 32556143; PMCID: PMC7337663.

31) Rosenberg ES, Dufort EM, Udo T, et al. Association of Treatment With Hydroxychloroquine or Azithromycin With In-Hospital Mortality in Patients With COVID- 19 in New York State. JAMA. 2020 Jun 23;323(24):2493–2502. doi: 10.1001/jama.2020.8630. PMID: 32392282; PMCID: PMC7215635.

32) Sbidian E, Josse J, Lemaitre G, et al. Hydroxychloroquine with or without azithromycin and in-hospital mortality or discharge in patients hospitalized for COVID-19 infection: a cohort study of 4,642 in-patients in France. medRxiv 2020.06.16.20132597; doi: https://doi.org/10.1101/2020.06.16.20132597

33) Singh S, Khan A, Chowdhry M, Chatterjee A. Outcomes of Hydroxychloroquine Treatment Among Hospitalized COVID-19 Patients in the United States- Real-World Evidence From a Federated Electronic Medical Record Network. medRxiv 2020.05.12.20099028; doi: https://doi.org/10.1101/2020.05.12.20099028

34) Skipper CP, Pastick KA, Engen NW, et al. Hydroxychloroquine in Nonhospitalized Adults With Early COVID-19: A Randomized Trial. Ann Intern Med. 2020 Jul 16:M20–4207. doi: 10.7326/M20-4207. Epub ahead of print. PMID: 32673060; PMCID: PMC7384270.

35) Sulaiman T, Mohana A, Alawdah L, et al. The Effect of Early Hydroxychloroquine-based Therapy in COVID-19 Patients in Ambulatory Care Settings: A Nationwide Prospective Cohort Study medRxiv 2020.09.09.20184143; doi: https://doi.org/10.1101/2020.09.09.20184143

36) Yu B, Li C, Chen P, et al. Low dose of hydroxychloroquine reduces fatality of critically ill patients with COVID-19. Sci China Life Sci. 2020 May 15:1–7. doi: 10.1007/s11427-020-1732-2. Epub ahead of print. Erratum in: Sci China Life Sci. 2020 Jun 18;: PMID: 32418114; PMCID: PMC7228868.

37) Matthew M Ippolito, Charles Flexner, Dose Optimization of Hydroxychloroquine for Coronavirus Infection 2019: Do Blood Concentrations Matter?, Clinical Infectious Diseases, ciaa691, https://doi.org/10.1093/cid/ciaa691

38) McGonagle D, Sharif K, O’Regan A, Bridgewood C. The Role of Cytokines including Interleukin-6 in COVID-19 induced Pneumonia and Macrophage Activation Syndrome-Like Disease. Autoimmun Rev. 2020;19(6):102537. doi:10.1016/j.autrev.2020.102537

39) Thachil J. The versatile heparin in COVID-19. J Thromb Haemost. 2020;18(5):1020–1022. doi:10.1111/jth.14821

40) Boulware DR, Pullen MF, Bangdiwala AS, et al. A Randomized Trial of Hydroxychloroquine as Postexposure Prophylaxis for Covid-19 [published online ahead of print, 2020 Jun 3]. N Engl J Med. 2020;NEJMoa2016638. doi:10.1056/NEJMoa2016638

41) Liu, J., Cao, R., Xu, M. et al. Hydroxychloroquine, a less toxic derivative of chloroquine, is effective in inhibiting SARS-CoV-2 infection in vitro. Cell Discov 6, 16 (2020). https://doi.org/10.1038/s41421-020-0156-0

42) Schmidt R. L., Jutz S., Goldhahn K., Witzeneder N., Gerner M. C., Trapin D., et al. (2017). Chloroquine inhibits human CD4(+) T-cell activation by AP-1 signaling modulation. Sci. Rep. 7:42191. 10.1038/srep42191

43) Jankelson L, Karam G, Becker ML, Chinitz LA, Tsai MC. QT prolongation, torsades de pointes, and sudden death with short courses of chloroquine or hydroxychloroquine as used in COVID-19: A systematic review. Heart Rhythm. 2020 Sep;17(9):1472–1479. doi: 10.1016/j.hrthm.2020.05.008. Epub 2020 May 11. PMID: 32438018; PMCID: PMC7211688.

44) Zakariya Kashour, Muhammad Riaz, Musa A Garbati, Oweida AlDosary, Haytham Tlayjeh, Dana Gerberi, M Hassan Murad, M Rizwan Sohail, Tarek Kashour, Imad M Tleyjeh, Efficacy of chloroquine or hydroxychloroquine in COVID-19 patients: a systematic review and meta-analysis, Journal of Antimicrobial Chemotherapy, dkaa403, https://doi.org/10.1093/jac/dkaa403

